# Facilitating the Virtual Exercise Games for Youth with Type 1 Diabetes (ExerT1D) Peer Intervention: Protocol Development and Feasibility

**DOI:** 10.1101/2024.07.03.24309308

**Authors:** Garrett I. Ash, Soohyun Nam, Selene S. Mak, Matthew Stults-Kolehmainen, Adrian D. Haughton, Carolyn Turek, Julien S. Baker, Kimberly Hieftje, Asher Marks, Annette Chmielewski, Michael Shelver, Elizabeth Considine, James L. Lukasik, Stuart A. Weinzimer, Laura M. Nally

## Abstract

**Aims:** Adolescents with type 1 diabetes (T1D) face several unique challenges to engaging in moderate-to-vigorous physical activity (MVPA), including uncertainty with diabetes self-management, risk of hypoglycemia, limited access to safe or supportive environments, and diabetes-specific stigma. Opportunities for T1D peer activities with T1D role model support are limited. To address this need, we tested iterative refinements of pilot Virtual Exercise Games for Youth with T1D (ExerT1D) for feasibility and acceptability.

**Materials and Methods:** Youth with T1D (ages 14-19) were recruited at a pediatric diabetes clinic and through social media. The program included 6 versions: study #1 (versions 1.1-1.4) included an active videogame, while study #2 (versions 2.1-2.2) included a virtual reality active videogame. All versions of the program included education about managing T1D with MVPA and objective habitual MVPA goal-setting guided by clinicians and young adult coaches living with T1D.

**Results:** Seventeen adolescents (median age 15.4 [IQR 14.6 - 16.4] years, 7 non-Hispanic white, 8 male, median HbA1c 8.1% [IQR 7.4%-11.1%]) enrolled. In total there were 260 person-sessions offered over 130 person-weeks. 15 participants attended 83% of sessions. Sessions began with pre-MVPA safety briefing with glucose checks, followed by the MVPA session, and ended with debriefing, goal-setting, and role-playing skits involving educational points related to T1D management. Participants rated the program, comfort, clinicians, coaches, and group cohesion high/very high (4.2±0.5 to 4.8±0.3 out of 5). They also rated motivation for the videogame high (4.1±0.4 out of 5). Building T1D and MVPA self-management skills achieved an excellent rating at a majority of sessions in both versions (76%), as did peer interactions (69%) and enriched communication (56%) after the addition of immersive virtual reality in version #2. Transitions between virtual reality apps forced group delays of 19±6min per session and some individuals to exercise separately. Compared to baseline, glycemic metrics appeared to decrease over time (d= -1.12, 90% CI [-1.78, -0.48]). No participants experienced diabetic ketoacidosis, severe hypoglycemia, or injuries during the study period.

**Conclusions:** Overall, ExerT1D facilitated excellent peer support by engaging diverse youth with T1D in an MVPA program led by role models living with T1D.

## INTRODUCTION

The adolescent years present unique challenges for youth with type 1 diabetes (T1D). Only ∼20% of youth meet the recommended A1c level below 7% shown to correlate with long-term health.^1,2^ Moderate-to-vigorous physical activity (MVPA) can be leveraged to improve glycemia, weight, and cardiovascular risk factors^3^. In addition, MVPA has been demonstrated to improve overall body composition, muscle strength, cardiopulmonary fitness, mental health, diabetes-specific risks of glycemic outcomes and insulin resistance.^3^ MVPA sustained in adulthood, is associated with less fewer microvascular complications and mortality.^4^ Managing glycemia during MVPA requires more frequent glucose monitoring and adjustments to diet and insulin dosing, that are further complicated in adolescence by impaired executive function^5^ and puberty-specific hormones.^3,6–8^ Perceived barriers to physical activity reported by adolescents include the fear of glycemic destabilization^9^ and social stigma (i.e., “negative social judgments, stereotypes, prejudices”^10^) associated with disclosing an individual’s diagnosis when engaging in MVPA.^11^ Thus, it is warranted to pursue multifaceted interventions targeted at addressing these barriers.

Current pediatric T1D MVPA guidelines are available but not actively understood or prescribed by most clinicians.^12^ Additionally, there are limited data addressing feasibility and acceptability of studies supporting physical activity for youth with low baseline MVPA.^3^ Active videogames and virtual reality (VR) are potential strategies to promote MVPA among youth.^13,14^ Group activities incorporating active videogame and VR technology with T1D mentors provide an exciting new avenue to deliver components of a successful T1D self-management intervention:^15^ 1) behavior modification (goal setting, diabetes self-management); 2) psychosocial support (engagement with T1D peers and role models); and 3) medical guidance (real-time diabetes self-management advice). Herein we present six cohorts of 3-5 participants evaluating the feasibility, acceptability, and preliminary safety of iterative refinements of a home-based virtual intervention to promote MVPA among adolescents with T1D.

## METHODS

### Enrollment

Adolescents (14-19yr old, T1D ≥6mo) not achieving recommended MVPA targets (i.e., <4days/wk with 60+min self-reported^16^) were recruited from the Yale Children’s Diabetes Center, clinicaltrials.gov (NCT05163912, NCT05662826), and diabetes social media groups between December 23, 2021 and June 15, 2022 (study #1; versions #1.1-1.4) and January 12^th^ – March 2^nd^, 2023 (study #2; versions #2.1-2.2). Screening was conducted by telephone using the Physical Activity Readiness Questionnaire verified against medical record^17^. For any health conditions presenting special considerations for MVPA, additional documentation was requested from the primary care physician or sub-specialist for MVPA clearance (n=1, arthritis). Informed consent or parental permission/assent (<18yr) was completed and signed using REDCap eConsent over a HIPAA-compliant tele-video call (Zoom Video Communications, San Jose, CA). Data on demographics, insulin therapy, anthropometrics, MVPA,^16^ and sedentary time^18^ were collected by parental REDCap survey and verified against medical record. Screening and intake data for study #2 were also used as long-term follow-up for study #1. The research team mailed supplies to the participant and held a second tele-video call to confirm proper setup.

### Intervention Development

The intervention, based upon Social Cognitive Theory, integrated peer and role model interactions into home-based MVPA^19^: networked active videogame sessions and virtual discussions about experiences managing diabetes and MVPA. In study #2, we added VR as a delivery tool to enhance the drivers of adolescent development and motivation according to self-determination theory, which are: competence, autonomy, and relatedness^20–22^. VR enhances competence by providing a safe space to learn competence in specific activities like MVPA and self-monitoring with a controlled degree of distractions and barriers. VR enhances autonomy by allowing users to embody a self-designed avatar. Finally, VR enhances relatedness by allowing users to experience new unique environments for the first time as a group.

Study #1 participants fitted their home television monitor with a Nintendo Switch Entertainment System (Nintendo of America, Inc., Redmond, WA) including Ring Fit Adventure which alternates low-(stationary walking or jogging) with 60+ higher-intensity exercises that are dynamic (e.g., knee-lifts) or resistance-based (e.g., abdominal press) tracked by a leg-strap accelerometer and handheld ring with gyroscopic/accelerometer sensors. Success with exercises unlocks more advanced adaptations (e.g., Russian twist). They were also provided a yoga mat, poster with Borg rating of perceived exertion scale, Precision Xtra Blood Ketone meter and strips (Abbott Diabetes Care, Alameda, CA), and Fitbit Inspire 2 (Google, Menlo Park, CA). Sessions were livestreamed over Zoom (Figure 1).

**Figure 1.**
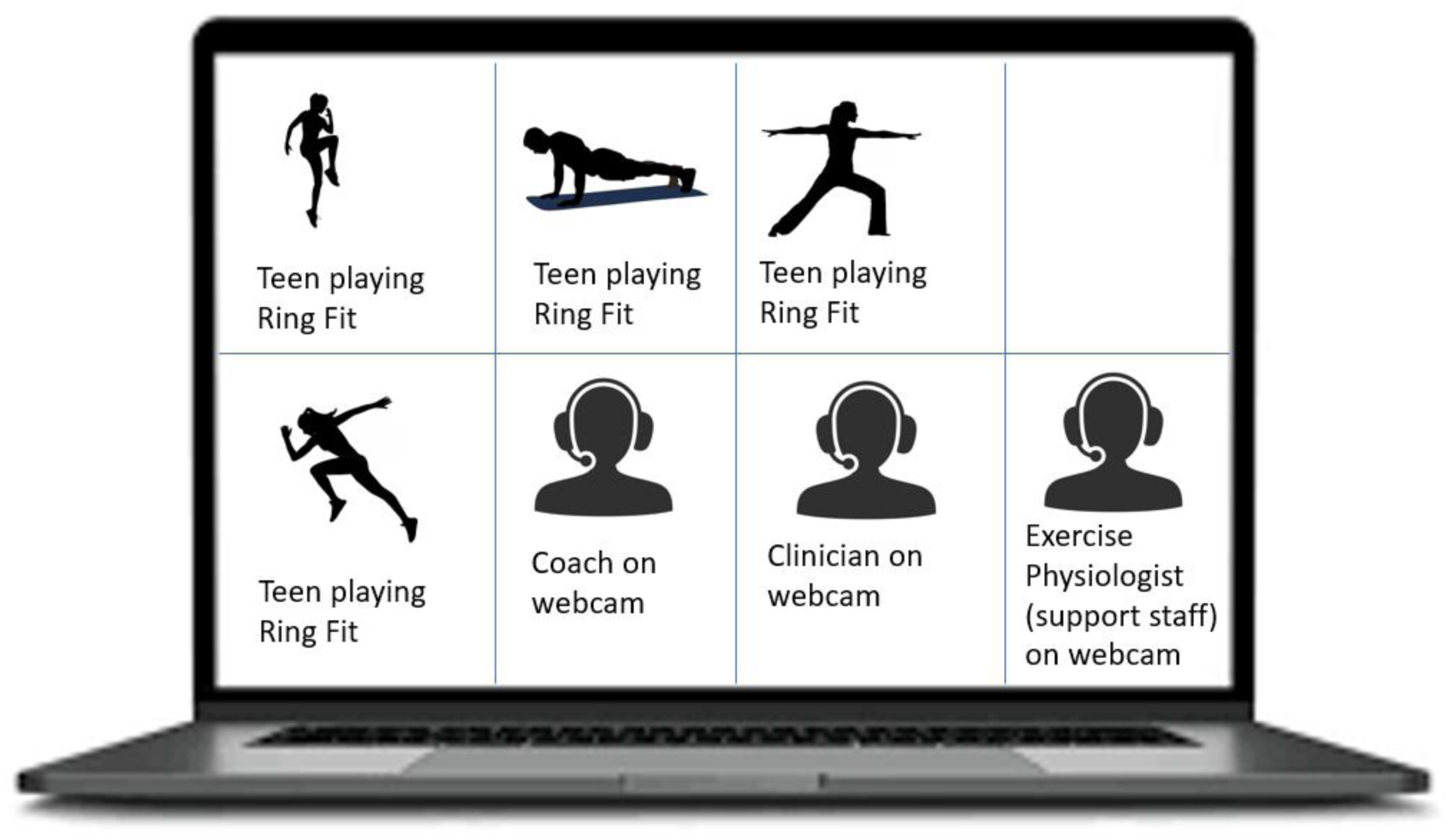
Study #1 virtual exercise session livestreamed over Zoom. Sessions were guided by a coach, and a clinician with expertise in T1D exercise management was present to advise diabetes-specific adjustments for exercise management.

Study #2 participants engaged in physical activity sessions using a virtual reality headset (Meta Quest 2 headset, Menlo Park, CA). Participants completed headset tutorials to adjust the lenses for proper focus, draw a guardian boundary to demarcate an area without obstacles or tripping hazards, and learn how to use the controllers. Participants also designed a unique Avatar to represent themselves during activity using Meta’s features (clothing, accessories, general appearance [e.g., skin tone, eye color, body shape], hairstyle, and voice modifications) and use two applications within the headsets. The first application, FitXR (Meta, Menlo Park, CA) facilitated group exercise classes using the avatars side-by-side to other participants to perform exercises guided by verbal and visual instructions from an automated avatar coach. Participants received individual and group scores within each activity. Following exercise activities, participants used VR headsets to meet in a virtual environment (Foretell Reality, Glimpse Group, New York, NY). Environments included a campfire with roasted marshmallow props, hot chocolate, and a guitar or a tropical beach with sea animals. These environments contain presence-enhancing features (spatial audio, hand and face tracking).

Four coaches living with T1D aged 20 to 26 years old (2 non-Hispanic White male, 1 non-Hispanic Black female, 1 non-Hispanic White female) with experience managing MVPA with T1D facilitated the sessions. The first coach was trained by reading a prototype manual and lesson guides prepared by the principal investigator with input from the multidisciplinary investigative team (pediatric endocrinologists, a pediatric psychologist, an exercise physiologist, an exercise telemedicine instructor, and behavioral interventionists). Then, the coach conducted two mock sessions with investigative team members as participants. The subsequent coaches were trained by reading the manual, observing recordings of prior sessions, and running mock sessions with the principal investigator and research assistants (RAs) as participants. All mock and live sessions were followed by feedback and refinements (c.f. Assessments below).

An RA facilitated intervention sessions and followed a source document to record the completion of each activity along with related diabetes-specific data (Appendix 1). The RA also helped participants troubleshoot technical difficulties with using the video games and equipment. A clinician (pediatric endocrinologist, certified diabetes care and education specialist) provided guidance about diabetes-specific exercise management according to published guidelines^3,23^ and an exercise physiologist monitored adherence to the intervention protocol (Appendix 1).

Participants were recruited in sequential cohorts for the 6-week (version 1) and 4 week (version 2) intervention with meetings twice weekly (Table S1) over Zoom. At the first session in each cohort, the clinician gave an overview of safe exercise practices related to T1D management based on national and international guidelines.^6,7,24^ Prior to activity, coaches confirmed that the participants had water, fast-acting carbohydrates to treat hypoglycemia, proper exercise shoes and clothing, and a 2.4m-by-2.4m unobstructed space for exercise. Participants communicated to the coach if they needed to leave the session for any reason, and the RA maintained a list of each participant’s physical location in case emergency services were needed.

### Diabetes Management

At each session, before and after exercise, teens reported their blood glucose or sensor glucose value, trend arrow, and insulin administration history (time and amount of last bolus and/or insulin-on-board from pump). Exercise was paused, and clinical guidance was provided if a glucose level was less than 70mg/dL and/or symptoms of hypoglycemia were reported. Ketone levels were checked if a glucose level exceeded 250mg/dL or there were clinical concerns for elevated ketones. Clinical guidance was provided when blood ketone levels were elevated ≥0.6mmol/L or urine ketones were moderate or large. On a weekly basis, the clinician reviewed continuous glucose monitor (CGM) and ketone data collected by questionnaire or upload (Keto-Mojo GK+, Napa, CA), identified any safety concerns (>4% time with hypoglycemia), and communicated these concerns with participants and parents.

For the purposes of coaching exercise, we tracked the proportion of each session spent in active movement, repetitions of each Ring Fit exercise or total FitXR target hits, Borg 6-20 rating of perceived exertion of each Ring Fit exercise, Borg 1-10 rating of the total session, and average Fitbit heart rate.

Following the 20-40min exercise activity, the last 20-30min of each session was spent on discussion over Zoom (version 1) or in virtual environments. Week 1 sessions were dedicated to creating individual fitness-related goals following the SMART (Specific, Measurable, Attainable, Relevant, Timely) principle. In addition, participants in each cohort formulated a group goal of cumulative achievements (e.g., group steps) to promote a cooperative rather than competitive approach.^25^ Goal progress was checked and discussed each week, including the provision of MVPA and T1D management information that helped support the specific goals. During middle week sessions, coaches engaged teens to create role-playing skits on Zoom that involved educational points related to T1D management. During the final sessions, the coaches led a discussion and painting exercise allowing participants to express the relevance of the intervention to building and sustaining an overall active lifestyle (Figure 2).

**Figure 2.**
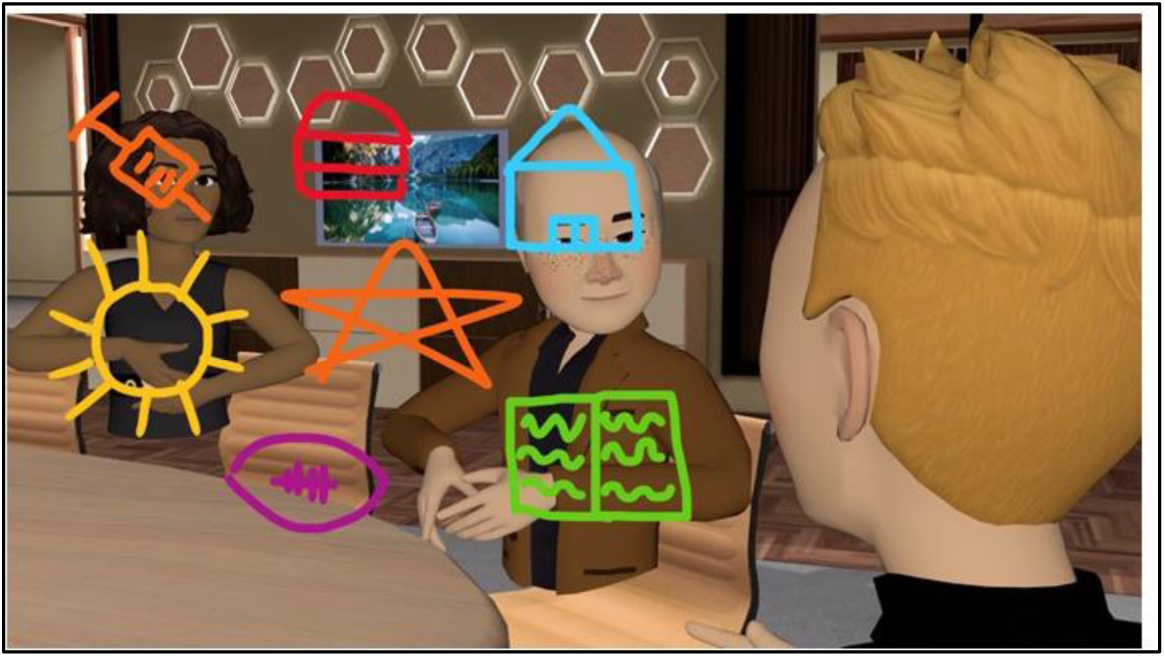
The version 2 virtual reality discussion environment included 3-dimensional painting where adolescents could express where their MVPA goals fit with their overall personal perspectives on MVPA, T1D, peers, family, other life topics, and their intersections. In the above example an adolescent represents themselves as a star that shines and dims with mood and life events, surrounded by diabetes (syringe), food managed with diabetes (hamburger), school, homework and projects (book), sports managed with diabetes (football), and being outside (runner). In version #1, similar discussions occurred verbally without visual expression.

### Assessments

Session recordings were double-coded by exercise physiologists or psychology RA to identify missed opportunities for practicing each core competency of a role model and peer intervention^26–31^ which were then discussed with instructors weekly to iteratively improve the intervention. The competencies were:

**1) Facilitating Peer Interactions.** We encouraged peer support with group discussions led by coaches, inviting each participant to share general life updates, discuss diabetes management challenges and successes, and physical activity goals.
**2) Practicing enriching communication.** Instructors and clinicians modeled empathy, resilience, and realistic optimism, validating challenges, empowering youth to manage their condition with confidence and self-compassion. The team encouraged conversations that were supportive, meaningful, and growth-oriented.
**3) Building T1D and MVPA Management Skills.** Clinicians led T1D self-management skill-building exercises, reviewing participants’ glucose changes during exercise shared in the physical activity session to augment glucose pattern recognition and problem solving, emphasizing that glucose levels during physical activity can vary based on recent insulin doses (“insulin on board”), carbohydrate intake and time of day. We also reviewed strategies to mitigate hypoglycemia and hyperglycemia during activity (e.g., reducing insulin prior to activity or setting an “activity” or “exercise” mode or “temp target” prior to and after physical activity).

At the end of the study, participants completed a follow-up retrospective survey of acceptability including the motivation for exergame play inventory (α=0.79),^32^ perceived cohesion scale (α=0.78-0.83),^33^ and a satisfaction survey from our past work^27^ to evaluate program components/strategies (5 items, α=0.72), personal comfort level (6 items, α=0.81), and interactions with instructors (12 items, α=0.95). Participants also rated the top 3 factors by importance.

### Feasibility Outcomes and Analytic Methods

Study outcomes and pre-specified thresholds included recruitment uptake (≥35%),^34^ attendance (≥75% attending ≥75% of sessions),^35^ biosensor wear-time (≥70%),^36^ survey completion (≥85%), fidelity ratings (≥70% excellent, ≥90% at least fair), and psychosocial perceptions of acceptability (≥3.0 out of 5). Variable distributions were described using visual inspection, Shapiro-Wilks test of normality, Levene’s test of equal variances, and appropriate effect size formulas to quantify temporal trends (SPSSv28, Chicago, IL). Inferential statistics such as ANOVA or comprehensive pairwise testing were not performed due to the small sample size. We refined the curriculum based on data after each cohort, ending the trial when no further refinements were found. This sample size (n=15) had 83% statistical power to detect any feasibility problems having ≥10% incidence according to Viecthbauer’s formula n = (ln[1-γ])/(ln[1-π]) where γ is power and π is minimum incidence of a feasibility problem to be detected.^37^

The intervention was unique in that it included both short-term behavioral reinforcement (e.g., peer encouragement of MVPA) and psychoeducation targeting long-term habit change (e.g., role-playing skits involving educational points). As such, we observed metrics not only during the 4-6-week interventions (e.g., CGM data) but also up to 1 year later (e.g., medical record HbA1c) in accord with standard practices for brief psychoeducational interventions (e.g.,^38,39^).

## RESULTS

### Participants

Seventeen participants referred by local clinicians, Facebook posts, and T1D community organizations were enrolled and completed follow-up (Figure S1), representing diverse geographic (8 states) and demographic characteristics: 8 female sex, 6 female gender, 2 non-binary gender, 4 public insurance, 2 with household incomes below the federal poverty line, and 7 household income <$60,000 (Table 1). Most participants had A1c levels above American Diabetes Association targets: 12 had an A1c of greater than 7.0%, and 5 had an A1c level of greater than 10.0%.

**Table 1.**
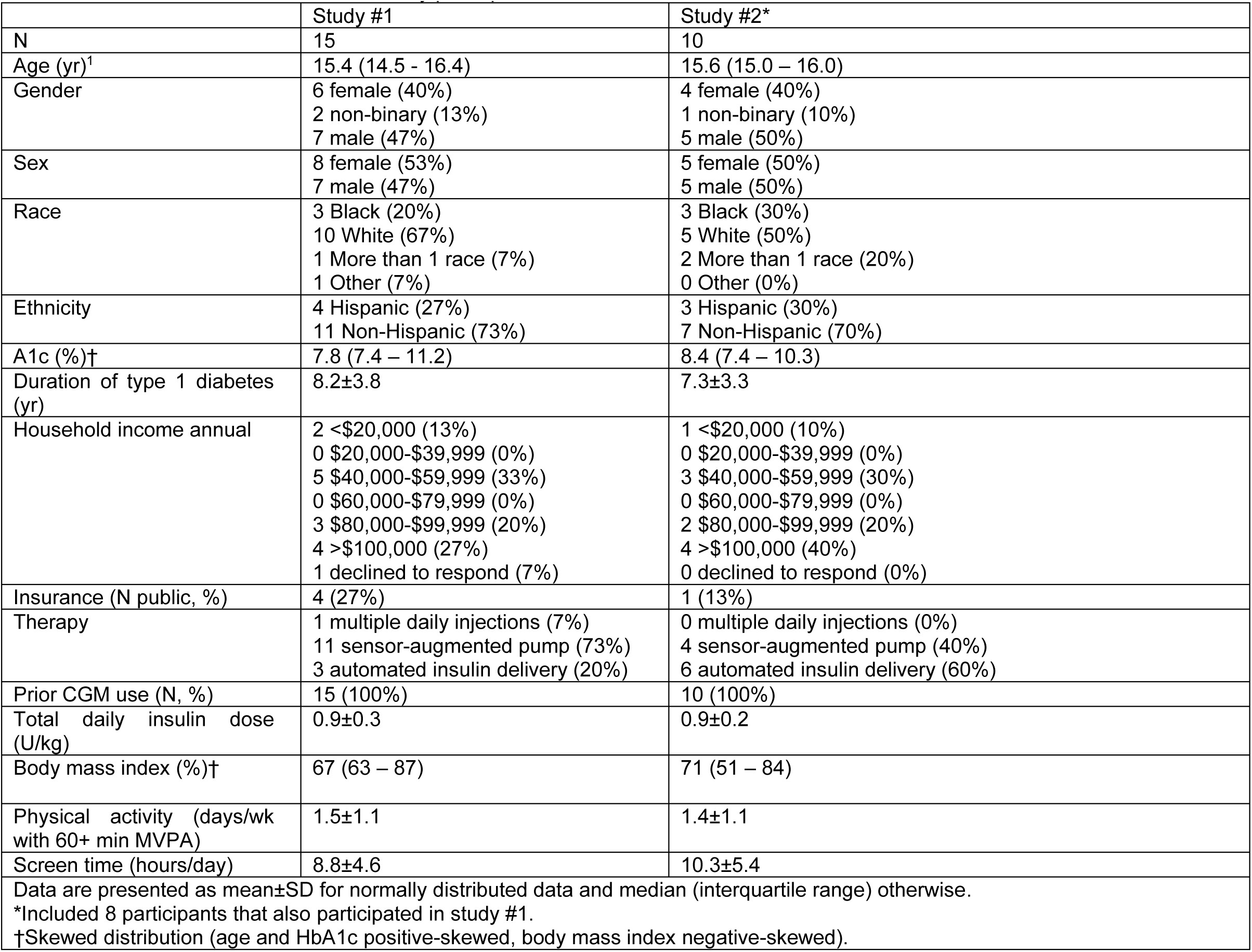
Baseline characteristics of the study participants.

|  | Study #1 | Study #2* |
| --- | --- | --- |
| N | 15 | 10 |
| Age (yr)<sup>†</sup> | 15.4 (14.5 - 16.4) | 15.6 (15.0 – 16.0) |
| Gender | 6 female (40%)<br>2 non-binary (13%)<br>7 male (47%) | 4 female (40%)<br>1 non-binary (10%)<br>5 male (50%) |
| Sex | 8 female (53%)<br>7 male (47%) | 5 female (50%)<br>5 male (50%) |
| Race | 3 Black (20%)<br>10 White (67%)<br>1 More than 1 race (7%)<br>1 Other (7%) | 3 Black (30%)<br>5 White (50%)<br>2 More than 1 race (20%)<br>0 Other (0%) |
| Ethnicity | 4 Hispanic (27%)<br>11 Non-Hispanic (73%) | 3 Hispanic (30%)<br>7 Non-Hispanic (70%) |
| A1c (%)<sup>†</sup> | 7.8 (7.4 – 11.2) | 8.4 (7.4 – 10.3) |
| Duration of type 1 diabetes (yr) | 8.2±3.8 | 7.3±3.3 |
| Household income annual | 2 <\$20,000 (13%)<br>0 \$20,000-\$39,999 (0%)<br>5 \$40,000-\$59,999 (33%)<br>0 \$60,000-\$79,999 (0%)<br>3 \$80,000-\$99,999 (20%)<br>4 >\$100,000 (27%)<br>1 declined to respond (7%) | 1 <\$20,000 (10%)<br>0 \$20,000-\$39,999 (0%)<br>3 \$40,000-\$59,999 (30%)<br>0 \$60,000-\$79,999 (0%)<br>2 \$80,000-\$99,999 (20%)<br>4 >\$100,000 (40%)<br>0 declined to respond (0%) |
| Insurance (N public, %) | 4 (27%) | 1 (13%) |
| Therapy | 1 multiple daily injections (7%)<br>11 sensor-augmented pump (73%)<br>3 automated insulin delivery (20%) | 0 multiple daily injections (0%)<br>4 sensor-augmented pump (40%)<br>6 automated insulin delivery (60%) |
| Prior CGM use (N, %) | 15 (100%) | 10 (100%) |
| Total daily insulin dose (U/kg) | 0.9±0.3 | 0.9±0.2 |
| Body mass index (%)<sup>†</sup> | 67 (63 – 87) | 71 (51 – 84) |
| Physical activity (days/wk with 60+ min MVPA) | 1.5±1.1 | 1.4±1.1 |
| Screen time (hours/day) | 8.8±4.6 | 10.3±5.4 |
| Data are presented as mean±SD for normally distributed data and median (interquartile range) otherwise. |  |  |
| *Included 8 participants that also participated in study #1. |  |  |
| †Skewed distribution (age and HbA1c positive-skewed, body mass index negative-skewed). |  |  |

## Intervention Feasibility

### Attendance and Adherence

Fifteen participants attended ≥80% of sessions, while the remaining two attended 50%-60% of the sessions. Absences were due to family activities (7), school activities (7), negative mood (6), oversleeping (6), acute illness (4), or no answer was given (2). Most of these absences (27/32, 84%) occurred on Saturdays, the few absences on Wednesdays being due to school activities (1), negative mood (3), or no answer was given (1).

All participants were using a CGM in their clinical care before the study (14 Dexcom G6, 3 Abbott Libre 2). Overall, weekly audits found CGM adherence was 73.6% in study #1 and 75.0% in study #2 of the possible 288 daily readings were captured across all study participants. Data completeness at the participant level was 79% (SD 25%) with 12 participants at ≥70%, 4 participants at 24%-54%, and 1 participant only sharing visual CGM data without a csv file (excluded from Tables 5-6).

Fitbit wear time on average met literature standards (70.5%, or 16.9 hours per day suggesting an average of 10-15 hours during waketime).^36^ Version 1.1 participants stopped wearing the Fitbit 2-4 weeks into the program, so we inserted a biweekly check of wear-time and troubleshooting after which this pattern did not repeat.

### Technical Barriers

In study 1, one participant was unable to participate in one session due to “dead” RingFit controller batteries (rated as moderate-high disruption). This participant was also unable to communicate verbally in two other sessions due to phone audio interruption (though could communicate by text chat). Two other participants had brief Zoom connectivity interruptions on 3 occasions respectively (reported as mild disruption).

In study 2, there were 15 malfunctions of the Oculus system (8 microphone, 3 speakers, 2 guardian boundary misplacement, 2 passcode), 9 with FitXR (5 times full class did not launch, 4 times individual participant ejected from class), and 5 with Foretell Reality Rooms (4 times individual participant locked out or ejected, 1 time no audio in room). FitXR was used in 12 of the 16 sessions and Foretell Reality was used in all 16 sessions. All issues resolved after restarting the app and/or system. FitXR classes were not started until all individuals could join the class without malfunctions, causing a delay of 19 (SD=6) minutes per session. When malfunctions occurred for individual participants mid-session, activities were paused unless rejoining was impossible. In these cases, a backup coach joined the participant in a separate FitXR class (4 instances) or group discussion (1 instance). Most malfunctions (80%) occurred during weeks 3-4 of study 2 before Meta updated its firmware (v50), when other users nationally reported similar issues.

### Fidelity

In version #1, intervention evaluators rated coach competencies highest for building T1D self-management and MVPA management skills followed by enriching communication and facilitating peer interactions at the group sessions (Table 2). Most competencies trended upward for each successive cohort except for cohort #4. Cohort #3 was the largest size and most balanced with respect to race & ethnicity, income, HbA1c levels, and recruitment source (Table 3). Cohort #4 had lowest size plus lowest attendance among those enrolled. One coach covered the first two cohorts and another coach covered the last two, both achieved similar average scores on most competencies. In version #2, facilitating peer interaction ratings increased and reached similar rating as building skills. Practicing enriching communication also increased, to a lesser extent and building skills remained high.

**Figure 3.**
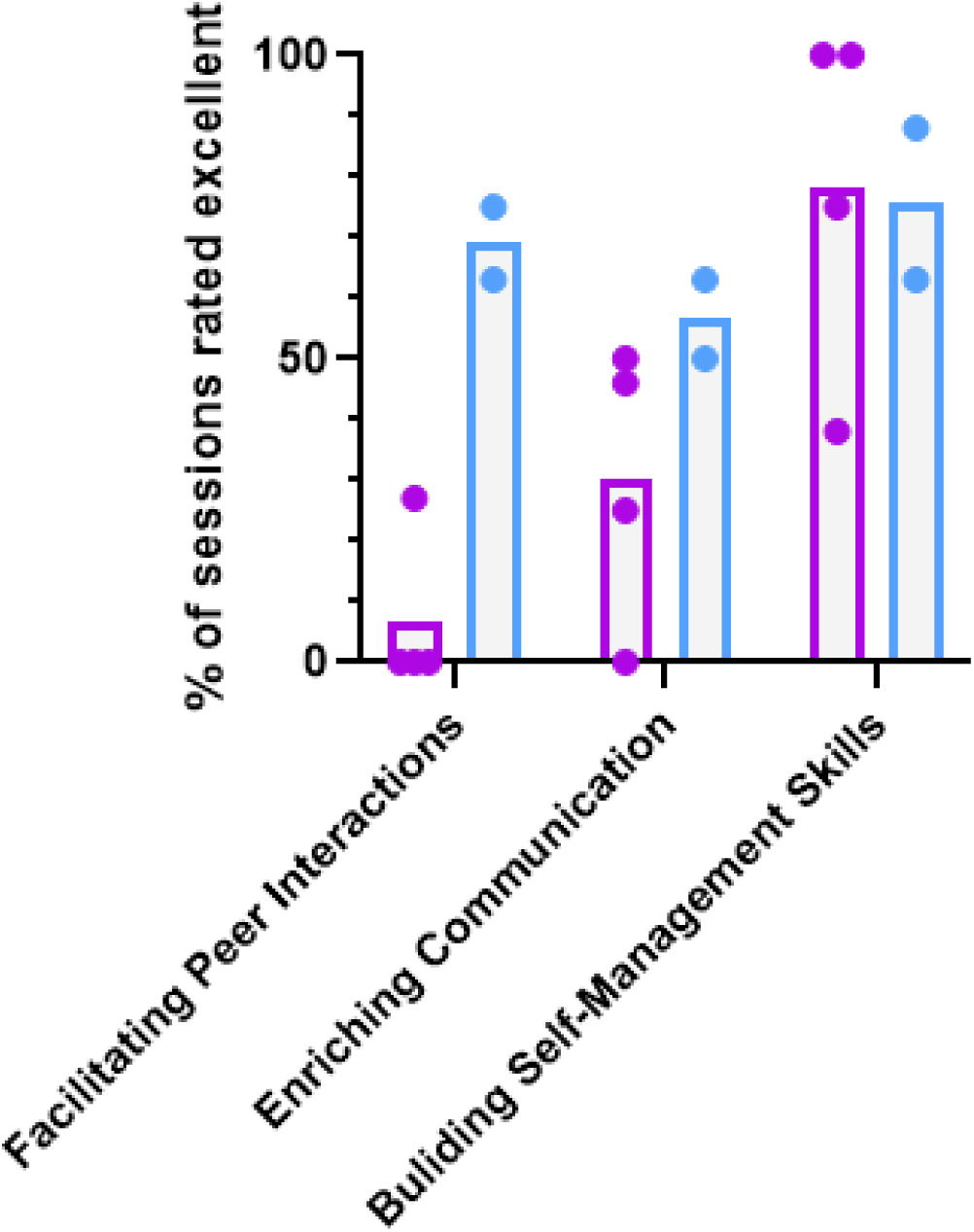
Competency ratings. Dots represent cohorts and are color-coded by study. Purple is study #1 (versions 1.1-1.4; 2-dimensional Ring Fit game and discussion by Zoom shown in methods Figure 1). Blue is study #2 (versions 2.1-2.2; virtual reality immersive environment shown in methods Figure 2).

**Table 2.** Fidelity, Satisfaction, Perceived Cohesion, and Exergaming Motivation Overall and by Cohort.

|  | **Instructor Competencies (% of sessions rated excellent)** |  |  | **Psychosocial Perceptions mean±SD (% rated ≥4 out of 5)** |  |  |  | **Exergaming Motivation mean±SD (% rated ≥4 out of 5)** |
| --- | --- | --- | --- | --- | --- | --- | --- | --- |
| **Version** | **Facilitating Peer Interactions at the Group Sessions** | **Enriching Communication at the Group Sessions** | **Building T1D and MVPA Management Skills** | **Interactions with Clinicians and Coaches** | **Group Cohesion (Perceived)** | **Program Components and Strategies** | **Comfort** | **Exergaming Motivation** |
| #1.1 (n=3) | 0 | 25 | 75 | 4.6±0.7 (67%) | 4.2±0.9 (67%) | 4.1±0.2 (100%) | 3.9±0.9 (67%) | 3.8±0.4 (33%) |
| #1.2 (n=4) | 27 | 46 | 100 | 4.9±0.0 (100%) | 4.6±0.4 (100%) | 4.5±0.3 (100%) | 4.5±0.1 (100%) | 4.1±0.2 (75%) |
| #1.3 (n=5) | 0 | 50 | 100 | 4.9±0.2 (100%) | 4.2±0.4 (80%) | 4.3±0.6 (60%) | 4.2±0.4 (80%) | 3.9±0.2 (60%) |
| #1.4 (n=3) | 0 | 0 | 38 | 4.8±0.3 (67%) | 4.7±0.2 (100%) | 4.3±0.7 (33%) | 4.3±0.6 (33%) | 4.6±0.2 (100%) |
| **Study #1 Total (n=15)** | **8** | **29** | **78** | **4.8±0.3 (93%)** | **4.4±0.5 (87%)** | **4.3±0.5 (73%)** | **4.2±0.5 (80%)** | **4.1±0.4 (60%)** |
| #2.1 (n=5) | 75 | 50 | 63 | 4.7±0.4 (100%) | 4.3±0.5 (80%) | 4.6±0.3 (100%) | 4.7±0.4 (100%) | 3.8±0.5 (40%) |
| #2.2 (n=5) | 63 | 63 | 88 | 4.5±0.5 (100%) | 4.4±0.8 (80%) | 3.8±0.2 (40%) | 3.7±0.6 (40%) | 3.9±0.4 (60%) |
| **Study #2 Total (n=10)** | **69** | **56** | **75** | **4.6±0.4 (100%)** | **4.3±0.6 (80%)** | **4.2±0.5 (70%)** | **4.1±0.7 (60%)** | **3.8±0.4 (50%)** |

Version 1.4 took one week longer than expected to recruit the minimum 3 participants. In fairness to the 2 participants who signed up on the preset schedule, we offered them 2 sessions during the week of delay which were credited as extra sessions.

**Table 3.** Cohort Characteristics.

| Cohort | Race/ Ethnicity | Gender | Age | Household Income (annual) | A1c | Recruitment | Attendance (Mean %)<sup>1</sup> |
| --- | --- | --- | --- | --- | --- | --- | --- |
| #1.1<br>(n=3) | 2 HispWhite<br>1 NHispWhite | 1 NB<br>2 M | 1, 14-15yr<br>2, 16-17yr | 1 under \$20k<br>2 under \$60k | 3 over 7.0%<br>1 over 10.0% | 3 from clinic<br>0 from social media | 75 |
| #1.2<br>(n=4) | 4 NHispWhite | 4 M | 3, 14-15yr<br>1, 16-17yr | 0 under \$20k<br>0 under \$60k | 2 over 7.0% | 0 from clinic<br>4 from social media | 92 |
| #1.3<br>(n=5) | 3 NHispBlack<br>1 More than 1 race<br>1 NHispWhite | 1 NB<br>3 F<br>1 M | 5, 14-15yr | 0 under \$20k<br>3 under \$60k<br>1 not reported | 5 over 7.0%<br>2 over 10.0% | 2 from clinic<br>3 from social media | 95 |
| #1.4<br>(n=3) | 2 HispWhite<br>1 NHispWhite | 3 F | 1 14-15yr<br>1, 16-17yr<br>1, 18-19yr | 1 under \$20k<br>2 under \$60k | 2 over 10.0% | 2 from clinic<br>1 from social media | 72 |
| #2.1 | 3 NHispBlack<br>2 More than 1 race | 3 F<br>2 M | 4, 14-15yr<br>1, 16-17yr | 0 under \$20k<br>3 under \$60k | 5 over 7.0%<br>2 over 10.0% | 2 from clinic<br>3 from social media | 90 |
| #2.2 | 2 HispWhite<br>3 NHispWhite | 1 F<br>3 M<br>1 NB | 4, 14-15yr<br>1, 16-17yr | 1 under \$20k<br>1 under \$60k | 4 over 7.0%<br>1 over 10.0% | 2 from clinic<br>3 from social media | 83 |

HispWhite, Hispanic White. NHispWhite, Non-Hispanic White. NHispBlack, Non-Hispanic Black. NB, non-binary. M, male. F, Female.
<sup>1</sup>Cohort #4 took one week longer than expected to recruit the minimum 3 participants. In fairness to the 2 participants who signed up on the preset schedule, we offered them 2 sessions during the week of delay which were credited as extra sessions.

### Intervention Acceptability

Participant satisfaction ratings in both versions were highest for interactions with clinicians and coaches, followed by perceived group cohesion, program components and strategies, comfort, and lastly, active videogame motivation (Table 2). The clinicians and coaches component was also most frequently selected as a top-1 or top-3 (72%, 64%).

### Intervention Safety and Monitoring

No participants experienced diabetic ketoacidosis, severe hypoglycemia, or other study-related adverse events during the study period. Two participants developed elevated ketone levels prior to exercise (1.1, 1.4 mmol/L) due to an overnight pump malfunction restricting their prior insulin, that resolved after insulin administration. One participant deferred exercise due to feeling unwell as a result of having a high sensor glucose level (400mg/dL) without elevated ketones. Three participants reported hypoglycemia symptoms during a session that resolved with carbohydrate treatment and three had sensor glucose levels <70mg/dL less than 1 hour after a session that participants resolved independently without staff guidance.

Weekly clinician reviews of CGM data resulted in 5 participants being referred to their healthcare providers for insulin dose adjustments to address elevated percentages of low sensor glucose values. Topics related to avoiding hypoglycemia after exercise were discussed in the group lesson, with participants directly.

Heart rate data were captured for 129/156 person-sessions (83%), with 15 out of 17 participants having at least one session. Among them, one averaged in the vigorous range of 70%-90% age-predicted maximum, 13 averaged in the moderate range of 55%-70%, and one averaged below the moderate range. The average participant heart rate across sessions was 61.4% ± 5.2% of maximum playing Ring Fit and 57.7% ± 6.0% playing FitXR.

### Intervention Acceptability

Ratings of specific components were the highest for activities as follows: group 3-D painting, seeing personal and peers’ scores for hitting boxing targets during MVPA, freely exploring the virtual reality environments, and exercising with other teens with T1D. The lowest-rated activities were those either administered in the non-avatar Zoom audio-chat (icebreaker questions, blood sugar checks) or automated videogame characters not part of the peer group (FitXR automated coach) (Table 4).

**Table 4.** Ranking of satisfaction of specific study #2 activities (n=10). Not done for study #1.

|  |  |
| --- | --- |
| **Component** | **Mean±SD (% rated ≥4 out of 5)** |
| Group 3D painting | 4.9±0.3 (100%) |
| Seeing individual score | 4.6±0.5 (100%) |
| Seeing everyone's scores | 4.4±0.7 (90%) |
| Freely exploring virtual environments | 4.4±0.7 (90%) |
| Exercising with other teens with type 1 diabetes | 4.3±0.7 (90%) |
| Exercise Movements | 4.2±0.6 (90%) |
| Exercising with coach that has type 1 diabetes | 4.1±0.6 (90%) |
| Diabetes skit | 4.1±1.0 (80%) |
| Talking about individual physical goals | 4.1±1.0 (80%) |
| Discussing exercise management strategies with clinicians and coaches | 4.1±0.7 (80%) |
| Talking about group physical activity goals | 4.1±0.7 (80%) |
| Icebreaker questions | 4.0±0.9 (80%) |
| Checking blood sugars and reporting them to clinicians and coaches | 4.0±0.9 (67%)† |
| Automated Exercise Coach | 3.7±0.7 (60%) |

†One participant declined to answer.

Participants set individual and group goals related to achieving Fitbit step counts, playing a sport, recreational exercise, or playing Ring Fit outside of group sessions, or managing diabetes (Table S2) with 28% success on individual goals and 50% success on group goals.

Average steps in study #1 for those with Fitbit step goals were 8274±3734 per day and for the total sample were 7178±4013 per day. Average steps in study #2 for the participant with a Fitbit step goal was 6,034 per day and for the total sample was 6467±2998 per day. Each skit successfully integrated 3-5 educational points (Table S3).

### Ancillary Analysis of Glycemic Outcomes

Between the two timepoints that HbA1c was available for the same participants - Study #1 baseline and follow-up (n=15) - it was on average lowered (8.9% vs 8.3%) but equally positive-skewed (Levene’s F_1,2_=0.458) (Figure 4). Such effect is best standardized as the Wilcoxon Signed Rank test index W= -1.905, equal to biserial r= -0.49 or Cohen’s d= -1.12 (90% CI [-1.78, -0.48]) (i.e., probably a medium or large effect).^40^ Total daily insulin showed no indication of increasing (0.88±0.27 to 0.83±0.24 U/kg/day) and participants upgrading to a closed-loop system during follow-up (n=7) had the same average HbA1c change as the full cohort. Body mass index percentile remained negative-skewed and showed no indication of change (median 67 vs 66, mean 75 vs 75, IQR 63-87 vs 53-86).

**Figure 4.**
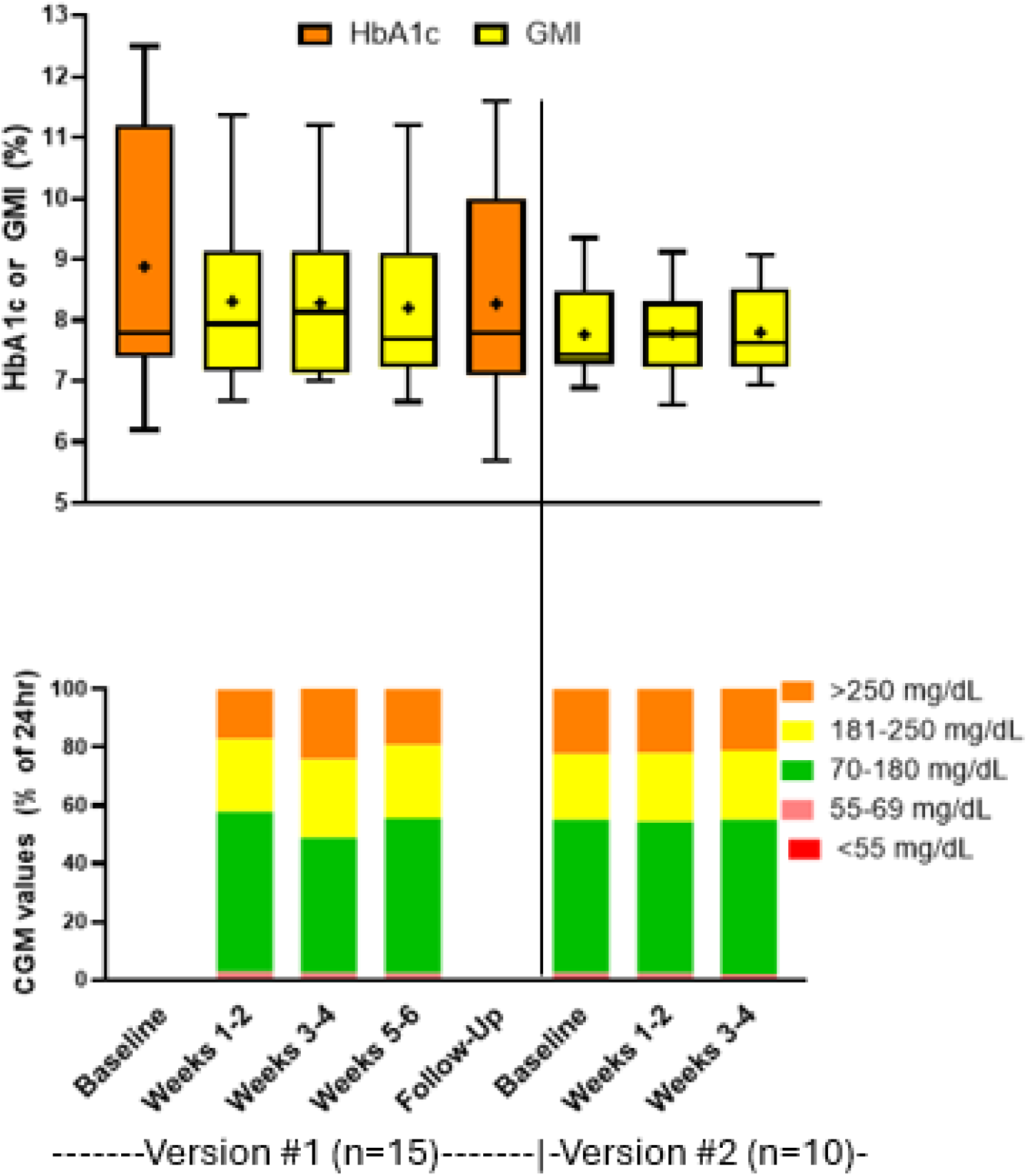
Glycemic metrics over time. **Top.** Metrics indicating average glucose. Version #1 baseline is laboratory HbA1c taken an average of 7 weeks (range 1-14) before intervention start, extracted from medical record. Version #1 follow-up is laboratory HbA1c taken an average of 45 weeks (range 19-77) after intervention start, extracted from medical record. Missing values (1 participant at weeks 1-6 due to not sharing downloadable CGM data, 2 participants at follow-up due to not sharing long-term access to medical record) were imputed by intention-to-treat, last observation carried forward. Other boxes are glucose management indicator (estimation of HbA1c based on CGM). Dash through box indicates median, plus sign indicates mean, box indicates interquartile range, whiskers indicate minimum and maximum. **Bottom.** Metrics indicating time in glucose ranges. Data completeness at the participant level was 79% (SD 25%) with 12 participants at ≥70%, 4 participants at 24%-54%, and 1 participant only sharing visual CGM data without a csv file (excluded). The 1 participant who did not share downloadable CGM data was excluded. Bars are omitted from timepoints where ≥5 participants had no shared data.

During the interventions, CGM clinical targets were met for time in hypoglycemia level 1 (1.8%-2.2% vs target ≤4%) and level 2 (0.5%-0.7% vs target ≤1%), but not hyperglycemia (42%-51% vs target ≤25%) or in target range (46%-55% vs target ≥70%) (Figure 4). Full CGM summary metrics are given in tables S4 and S5.

## DISCUSSION

The primary finding of the study was that the virtual, compared to in-person protocol,^26^ greatly increased feasibility while maintaining excellent safety standards and opportunities to interact with peers, clinicians, and young adult role model coaches. Compared to our in-person group, we doubled recruitment uptake proportion from 16% to 35%, which is comparable to 37% seen in personalized MVPA prescription for T1D that did not require fitting a group schedule.^34^ We also expanded from clinic to social media venues. In addition, we improved the attendance from 56% to 85%-90%, while also doubling the frequency of sessions from once to twice per week. A glucose monitoring protocol was followed with no adverse events and occurrence of MVPA-induced hypoglycemia no more frequently than typical daily living. The program was also highly cost-efficient. Social media advertisements utilized community postings rather than paid advertisements, and virtual technology was all within the scope of devices normally owned by diverse families of teens with T1D. Videogame technology is popular among teens and substantially more so among those from lower-income families.^41^

The high acceptability of the in-person protocol^27^ was also maintained based on mean evaluation scores, though unbalanced across subscales. Building T1D and MVPA self-management skills achieved an excellent rating at a majority of sessions in both versions (76%), as did peer interactions (69%) and practicing enriching communication (56%) after the addition of immersive virtual reality in version #2. Perceived group cohesion was also rated highly by the teens, reflecting some combination of perceived cohesion to the instructors and other teens. Program structure and comfort were rated less highly than clinician and coach interactions and perceived group cohesion, though nonetheless received ratings of 4+ out of 5 and were likely essential to the program’s feasibility.

Prior studies supporting MVPA for adolescents with T1D provided self-management education and decision guidance^34,42–44^ and only one included personal support, which came from key family members such as a parent.^34^ In real-world contexts, 91% of adolescents with T1D report that their parents discourage MVPA^9^ and that teachers and coaches have limited T1D knowledge.^45^ Therefore, interventions aimed at improving health outcomes need to be introduced during the transition period of adolescence when youth are developing the skills they will need as adults to manage T1D.^26,27^

Support from peers and peer mentors is rated as important by adolescents with T1D^46,47^ and associated with increased diabetes self-care.^48^ However, the incorporation of peers and role models is in nascent stages^28,49^ and mostly focused on T1D-specific behaviors rather than those important for general health such as MVPA. Such restriction of peer support to T1D-specific behaviors has been negatively associated with diabetes self-care,^48^ similar to the “nagging” perceived from parents.^50^ It may also be less engaging, as these prior peer interventions have been almost entirely restricted to asynchronous communications without physical activities.^29,30^ When synchronous chats have been added, just 39%-48% of participants attended one or more sessions.^30,49^ Overall, the present study shows the promise of peer group MVPA as a novel strategy both for supporting MVPA and generally engaging adolescents with T1D.

A major strength of the study was the enrollment of a diverse sample of adolescents in greatest need of support for MVPA and T1D self-management (lower income, people of color, elevated HbA1c).^51^ This study had no A1c upper limit, thus incorporating a group of youth with T1D at high risk of dysglycemia that has been largely excluded from research studies. Intervention attendance was excellent and overcame logistical barriers related to alignment of group schedules, availability of MVPA supplies, timing MVPA with insulin and diet, and provision of real-time guidance. Given the focus on feasible uptake of technology and inclusion of youth with all HbA1c levels, we enrolled participants even if they were wearing their CGM inconsistently at enrollment, which meant glycemic outcomes could only be assessed by combining HbA1c and GMI values in an ancillary analysis. HbA1c and GMI can be discordant due to the timeframe they reflect and non-glycemic factors that can impact HbA1c.^52^ Nonetheless, glycemic reductions were seen during and after the intervention. These findings were encouraging, especially in light of the elevated HbA1c levels in the cohort at baseline, as these youth are largely excluded from research.

We acknowledge some limitations of the intervention. Coaches were trained in group facilitation strategies derived from our expert planning group discussions and two pilot sessions, but for future studies we will utilize an accredited group facilitation course. Second, Fitbit measurements of heart rate may be less accurate for those with darker skin tone.^53^ However, Fitbit was mainly used for step-counting which does not have this limitation, and heart rate was only used for an assessment of convergent validity with the validated videogame.^54^ That the study was limited to those who were able to afford Internet access at home and time to attend the sessions does limit some generalizability and implementation. Finally, as a small single-group study design, we had limited ability to robustly determine efficacy outcomes.

## CONCLUSIONS

In summary, we report protocol development and feasibility of a virtual home intervention to promote MVPA among peer groups of adolescents with T1D. Successes included attraction of a diverse group, high attendance at the sessions, T1D self-management guidance that promoted safe MVPA, and meaningful connections to including shared experiences and skills in T1D self-management and coping. The challenge was technical delays especially transitions between virtual reality apps. Therefore, future trials testing efficacy and moderators warrant financial investment to integrate and interoperate the virtual reality apps. The advantages of our program were effectiveness at connecting teens to peers, clinicians, and young adult coaches in a way that was logistically feasible, time efficiency for the clinicians and coaches since they could interact with a group all at once, and sustainment of favorable MVPA and clinical metrics.

## DATA AVAILABILITY

Individual de-identified participant data (including data dictionaries) were shared. This includes individual participant data that underlies the results reported in any aspect of a published article (text, tables, figures, and appendices). Other documents that will be available include the study protocol, statistical analysis plan, informed consent form, and analysis code. The data will be available immediately following publication, with no end dates. The data will be shared with researchers who provide a methodologically sound proposal to achieve the aims of the approved proposal. The proposals should be directed to Dr. Garrett Ash at Yale University. To gain access, data requesters must sign a data access agreement.

## AUTHORSHIP CONFIRMATION/CONTRIBUTION STATEMENT

**GIA:** Conceptualization (lead), data curation (lead), formal analysis (lead), funding acquisition (lead), investigation (equal), methodology (equal), project administration (lead), resources (lead), software (support), supervision (lead), validation (equal), visualization (lead), writing – original draft (lead). **SN:** Conceptualization (equal); investigation (equal); methodology (equal); writing – review and editing (equal). **MSK:** Methodology (supporting); Writing – review and editing (equal). **ADH:** Investigation (equal); methodology (equal); project administration (equal); software (equal); writing, review, and editing (supporting). **CT:** Investigation (equal); methodology (equal); writing, review, and editing (equal). **AC:** Investigation (equal); methodology (equal); writing – review and editing (equal). **MS:** Investigation (equal); methodology (equal); writing, review, and editing (equal). **JSB:** Methodology (supporting); Writing – review and editing (equal). **SAW:** Conceptualization (equal); investigation (equal); methodology (equal); writing – review and editing (equal); writing – review and editing (equal). **LMN:** Conceptualization (equal), formal analysis (equal), investigation (equal), methodology (lead), software (lead), visualization (equal), writing – review and editing (lead).

## ACKNOWLEDGMENTS

The authors thank the families and participants that help made this research possible. We also acknowledge Sa’Ra Skipper and Juanita Montoya for their assistance with conducting the study. Study data were collected and managed using REDCap electronic data capture tools hosted at Yale University.

## Supplemental Material

**Figure S1.**
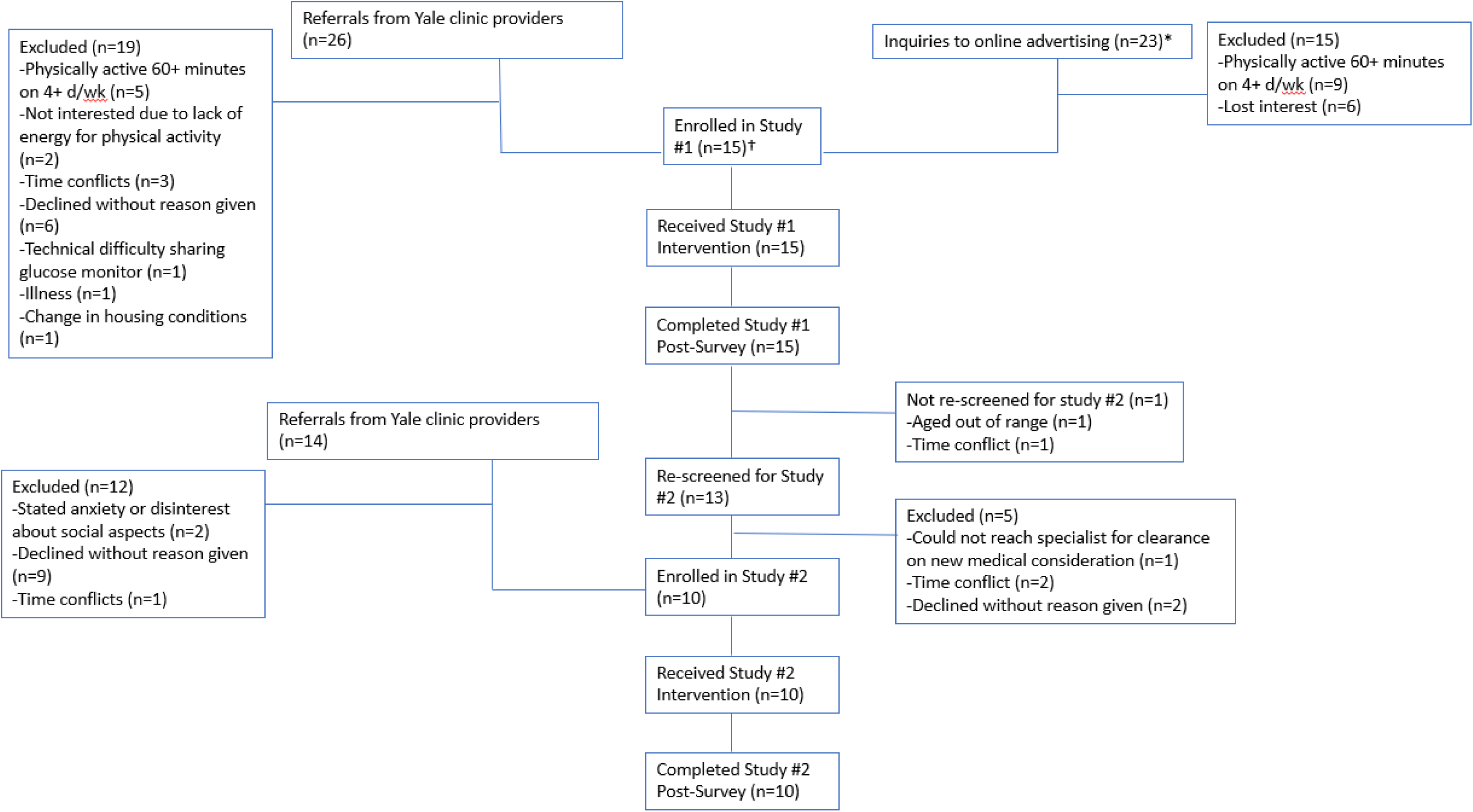
Enrollment flow-chart. *Online advertising consisted of Children with Diabetes posting the advertisement in three issues of their weekly newsletter (February 9^th^–February 23^rd^, 2022) as well as their Facebook page on February 15^th^. The Facebook post received 12 likes, 10 shares, and 23 email inquiries as listed. †One participant required clearance for a medical condition from a specialist (arthritis) prior to enrollment.

**Table S1.** Structure of intervention sessions.

| **Component** | **Saturdays Time of Day<sup>1</sup>** | **Wednesdays Time of Day** |
| --- | --- | --- |
| Icebreaker question | 10:00-10:05 | 19:00-19:05 |
| Sensor glucose check and guidance<sup>2</sup> | 10:05-10:20 | 19:05-19:20<sup>3</sup> |
| Exercise | 10:20-11:00 | 19:20-19:40<sup>3</sup> |
| Sensor glucose check and guidance | 11:00-11:05 | 19:40-19:45<sup>3</sup> |
| Discussion Activities | 11:05-11:30 | 19:45-20:00 |
| • MVPA goals (**week 1:** set goals; **weeks 2-5:** check progress and provide MVPA and T1D management information supporting goals; **week 6:** reflection on relationship to overall active lifestyle)<br>• Role-playing skits (**weeks 2-5:** planning and acting) |  |  |

<sup>1</sup>USA eastern time zone, which was the local time of all participants except 1 who was USA mountain time zone (2 hours earlier)
<sup>2</sup>Extended to 30 minutes at first session to give general overview of guidelines. This session had reduced exercise by 8 minutes and discussion activities by 5-10 minutes.
<sup>3</sup>Cohort #1 only had these components on Saturdays (i.e., Wednesday session only met 19:00-19:20). Added these components to Wednesdays per expressed interest of cohort #2.

**Table S2.** Individual Goals (n=15)

| **Goal Category** | **Number of participants** | **Specific Value** | **Achieved Goal** |
| --- | --- | --- | --- |
| Steps | 9 | School to home, 3d/week<br>School to home, every day<br>One mile around neighborhood, every day<br>4000 per day<br>7000 per day<br>8500 per day<br>10000 per day<br>12500 per day<br>13000 per every other day | 2 / 9 (22%) |
| Sport | 5 | Find a soccer group<br>Bike 26.2 miles<br>Run 8:00 mile<br>Run 6:35 mile<br>Run 7:30 pace for 5 miles<br>Dead lift 230 lbs<br>Push-up reps 20<br>Bike 30min per day<br>Pole vault reach minimum club competition clearance<br>Gym twice per week<br>Exercise 3 days per week<br>Run timed mile each week | 4 / 12 (33%) |
| Ring Fit | 3 | 20 minutes per day<br>500 reps of 1 specific movement<br>Reach world 9 | 1 / 3 (33%) |
| Diabetes management | 1 | 7.0% estimated A1c from CGM | 0 / 1 (0%) |
| Total | 15 |  | 7 / 25 (28%) |

**Table S3.** Structure of topics, roles, and competencies taught during skits for each cohort.

| Topic | Roles<sup>1</sup> | Educational Points and How Integrated |
| --- | --- | --- |
| Team sports and type 1 diabetes (T1D) | -Athlete with newly diagnosed with T1D<br>-Teammate living with T1D for several years<br>-Continuous glucose monitor with alerts<br>-Coach | -Speaking to coaches who are not familiar with T1D management around MVPA<br>-Peer mentoring<br>-Competitive sports and glycemic management |
| Zombie Apocalypse | -Zombie<br>-Zombie-killing engineer<br>-Teen newly diagnosed with T1D and knows nothing about what they need for T1D management during an apocalypse<br>-Teen with longer duration of T1D<br>-Dr. Rescue<br>-News host<br>-Continuous glucose monitor with alerts | -Essential supplies for T1D<br>-Prioritizing aspects of T1D management<br>-Operating continuous glucose monitors<br>-Impact of MVPA on glycemic management<br>-Impact of stress on glycemic management |
| Wilderness Excursion | -Lilo (Hawaiian girl)<br>-Stitch (helpful alien)<br>-Evil singing crab<br>-Battler of evil singing crab<br>-Diabuddy (diabetes buddy)<br>-Angel<br>-Narrator | -Same as Zombie Apocalypse |

<sup>1</sup>Self-chosen by interested participants, remaining ones assigned. MVPA, moderate-to-vigorous physical activity

**Table S4.** Summary of CGM for study 1. Reported as mean (SD). Normally distributed except where noted.

| Time Frame | Weeks 1-2 | Weeks 3-4 | Weeks 5-6 |
| --- | --- | --- | --- |
| Mean glucose (mg/dL) | 200.5 (53.1) | 199.2 (40.8) | 195.6 (41.9) |
| SD of mean glucose (mg/dL) | 73.6 (17.6) | 76.3 (18.6) | 73.8 (19.4) |
| CV (%) | 37.4 (6.9) | 38.7 (8.1) | 38.0 (4.5) |
| % time < 54 mg/dL<sup>1</sup> | 0.7 (1.0) | 0.8 (1.1) | 0.4 (0.6) |
| % time <70 mg/dL<sup>1</sup> | 2.8 (2.7) | 3.2 (4.4) | 2.7 (2.3) |
| % time 70-180 mg/dL | 46.2 (22.2) | 44.0 (18.6) | 48.6 (21.0) |
| % time >180 mg/dL | 51.0 (22.4) | 52.8 (19.4) | 48.5 (20.1) |
| % time > 250 mg/dL | 27.6 (24.0) | 27.5 (18.6) | 25.1 (20.0) |
| % time with sensor data | 74.3 (33.6) | 73.6 (31.8) | 72.8 (33.8) |

Abbreviations: CGM: continuous glucose monitor, SD: standard deviation, CV: coefficient of variation.
<sup>1</sup>Right-skewed. Specifically, more than one-third of participant-weeks had zero or negligible values: <0.2% for % time <54 mg/dL and/or <1.0% for % time <70 mg/dL.

**Table S5.** Summary of continuous glucose monitoring, insulin, and anthropometric data for study 2. Given as mean (SD).

| Time Frame | Weeks 1-2 | Weeks 3-4 |
| --- | --- | --- |
| Mean glucose (mg/dL) | 189.1 (33.1) | 186.0 (28.8) |
| SD of mean glucose (mg/dL) | 74.3 (18.0) | 73.5 (15.9) |
| CV (%) | 39.0 (4.0) | 39.3 (3.4) |
| % time < 54 mg/dL<sup>1</sup> | 0.7 (1.3) | 0.7 (1.5) |
| % time <70 mg/dL<sup>1</sup> | 2.4 (2.1) | 2.1 (2.2) |
| % time 70-180 mg/dL | 51.9 (16.7) | 53.0 (16.2) |
| % time >180 mg/dL | 45.7 (16.2) | 44.9 (14.5) |
| % time > 250 mg/dL | 22.0 (15.2) | 21.2 (13.9) |
| % time with sensor data | 75.0 (30.5) | 75.0 (36.7) |

Abbreviations: SD: standard deviation, CV: coefficient of variation.
<sup>1</sup>Right-skewed. Specifically, more than one-third of participant-weeks had zero or negligible values: <0.2% for % time <54 mg/dL and/or <1.0% for % time <70 mg/dL.

